# Global, regional, and national levels and trends in burden of urticaria: a systematic analysis for the global burden of disease 2019 study

**DOI:** 10.1101/2023.09.15.23295635

**Authors:** Yuanchun Pu, Liyu He, Xiangyu Wang, Yaodong Zhang, Shidi Zhao, Jinhai Fan

**Affiliations:** Department of Urology, The First Affiliated Hospital of Xi’an Jiaotong University, Xi’an 710061, China; Department of Radiology, The First Affiliated Hospital of Xi’an Jiaotong University, Xi’an 710061, China; Department of Urology, The Second Affiliated Hospital of Xi’an Jiaotong University (Xibei Hospital); Cancer Center, The First Affiliated Hospital of Xi’an Jiaotong University, Xi’an 710061, China

## Abstract

**Background:** Urticaria causes a significant burden on individuals and society due to its pervasiveness. The aim of this study was to evaluate the burden of urticaria in different regions and nations by analyzing data from the Global Burden of Disease Study 2019 (GBD 2019) to provide a reference for healthcare policymakers.

**Methods:** Using the GBD 2019 database, this study analyzed incidence, prevalence, disability-adjusted life year (DALY), and corresponding ASRs and EAPC globally and in 204 countries and regions, and stratified the data by age, sex, and sociodemographic index (SDI).

**Results:** In 2019, the global incidence cases of urticaria increased to 114708912.2, the prevalence cases increased to 65139886.6, and the global DALY burden increased to 3898838.6. The distribution of the burden was markedly geographically heterogeneous. Burden indicators were generally higher in females than in males. There was a slight positive correlation between urticaria burden and regional SDI.

**Interpretation:** The burden of urticaria has been increasing globally from 1990 to 2019. This study identified multiple determinants that influence the burden of urticaria, such as geographic location and SDI. Despite the limitations of the GBD 2019 data, these discoveries provide a valuable resource for the development of future public health strategies aimed at reducing the burden of urticaria.

## 1 Introduction

Urticaria, also known as hives, is a transient dermatological phenomenon characterized by pruritic, erythematous, and elevated plaques, often resulting from the degranulation of mast cells in the superficial dermis releasing histamine, and usually resolves within 24 hours.(1–3) Another manifestation associated with urticaria is angioedema, involving deeper blood vessels, specifically small veins, and these two conditions can sometimes coexist.(1,3) Although clinical attention is important, the potential societal burden is also heavy. In a recent meta-analysis, it was demonstrated that chronic urticaria alone possesses a lifetime prevalence of 1.4%, excluding other types of urticaria.(4) There are also studies using the Chronic Urticaria Quality of Life Questionnaire (CU-Q2oL) and Dermatology Life Quality Index (DLQI) surveys which show that the symptoms of urticaria can have a significant impact on patients’ sleep and daily life, as well as the work performance and productivity.(5) In addition to the personal impact, urticaria also has a high social cost including medical resources and money.(6,7) And there also are some studies shows the incidence rates exhibit significant variance across different countries which means that the burden of urticaria is common and variable.(7–9)

Although previously published papers have made many contributions to the study of the urticaria burden, no research has comprehensively analyzed and discussed the global burden from different perspectives spanning the years 1990 to 2019.(10–12) Even In the recently published “Letter to the Editor” that focuses on the burden of urticaria, the analysis remains incomplete with respect to both the current burden and its trends, while also failing to consider the etiological factors of the burden. (13) In this study, we describe the incidence, prevalence, and Disability-Adjusted Life Years (DALY) and analyze the Estimated Annual Percentage Change (EAPC) and Age-Adjusted Percentage Change (AAPC) from 204 countries and territories to evaluate the burden of urticaria and explain its developing trends at global, regional, national and Socio-demographic Index (SDI) scales. Our ultimate goal is to increase policymakers’ attention to the urticaria burden and promote rational allocation of health resources.

## 2 Materials and methods

### 2.1 Data Source

The Global Burden of Disease (GBD) database is an open-access public database with data from Literature, Survey Data, and Claims Data subjected to age-sex stratification. The GBD data of urticaria were analyzed by the Dismod-MR 2.1 computational tool with the integration of study-level covariates (Claims data - 2000), aimed at getting the prevalence and incidence of urticaria, classified by location, year, age, and sex. At the same time, information collected from the Medical Expenditure Panel Survey was used for the meta-analysis of the proportion of urticaria cases classified as either mild or severe. These bifurcated data streams were then combined and allocated Disability Weights. By using the Comorbidity Corrections (COMO), the data can calculate the years lived with disability (YLDs), which were further used to estimate DALYs.(14)

The most recent GBD 2019 Data Resources is used in this study and obtained by the Global Health Data Exchange (GHDx) query tool (http://ghdx.healthdata.org/gbd-results-tool) (accessed on 26 September 2022).(15) We select the data conforming to ICD10 urticaria diagnostic criteria (ICD-10:50), with no additional inclusions or exclusions. Symptoms of urticaria are categorized into two levels, and both get the corresponding disability weights: Mild urticaria: (Severity level 1, DW (95%CI:0.027 0.015–0.042)); Severe urticaria: (Severity level 2, DW (95%CI:0.188(0.124–0.267)).(14) The data we obtained includes multiple epidemiological indicators of 23 age groups: including incidence, prevalence, and YLDs, DALYs, and metrics used for these indicators include number, rate, percent, and years. Gender is grouped into males, females, and both sexes. The geographical scope comprises 204 countries and regions, divided into 21 regions and 7 larger areas (WHO regions, World Bank Income Levels, and more). Instead of using the Human Development Index (HDI) in other studies, we choose the Socio-Demographic Index (SDI) to make more accurate comparisons of the indicators related to medical health.

### 2.2 Statistical Analysis

In this study, we calculate Age-Standardized Rate (ASR), EAPC, and AAPC. ASR is used to neutralize the influence of age-structural differences when comparing urticaria risks. The calculation formula is: 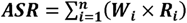 (*W_i_* is the proportion of the ***i^th^*** age group in the standard population; ***R_i_*** is the crude rate for the ***i^th^***age group in the observed population; ***n*** is the number of age group). EAPC is used to describe the trend of rate over a time period, and it’s calculated via a log-linear model: **ln(*y*) = *α* + *β* × calendar year + *∈***; (*y* is the annual rate; ***β*** shows the positive or negative trends of ASR; is the random error term) and EAPC is calculated as: **EAPC = (*e^β^* − 1) × 100**. Like EAPC, AAPC is also used to assess long-term trends but represents a weighted average of EAPCs over a given time span. It’s estimated through the Joinpoint model: 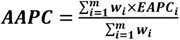 (*EAPC_i_* is the EAPC in the ***i^th^*** time; ***W_i_*** is the length of the ***i^th^*** time; ***m*** is the number of time segments). The p-value less than 0.05 was considered statistically significant.

Data analysis and data visualization were executed via R software (Version 4.1.2, https://www.r-project.org) and Joinpoint Trend Analysis Software (Versions 4.9.0.1, https://surveillance.cancer.gov/joinpoint/). The GBD study did not require patient informed consent and adhered to The Guidelines for Accurate and Transparent Health Estimates Reporting (GATHER) in population health research.

## 3 Results

### 3.1 Global urticaria burden

In 2019, there were 114708912.2 (95%UI=101309687.1-129286333.7) incidence cases globally with an age-standardized incidence rate (ASIR) equating to 1527.5 (95%UI=1346.2-1726.5) per 100,000 population. Meanwhile, the EAPC (0.022 (0.0176-0.0263)) and AAPC of incidence were both noted to be greater than 0, thereby showing an upward trend in the ASIR from the period of 1990 to 2019 (Table 1, Figure 1A).

**Figure 1.**
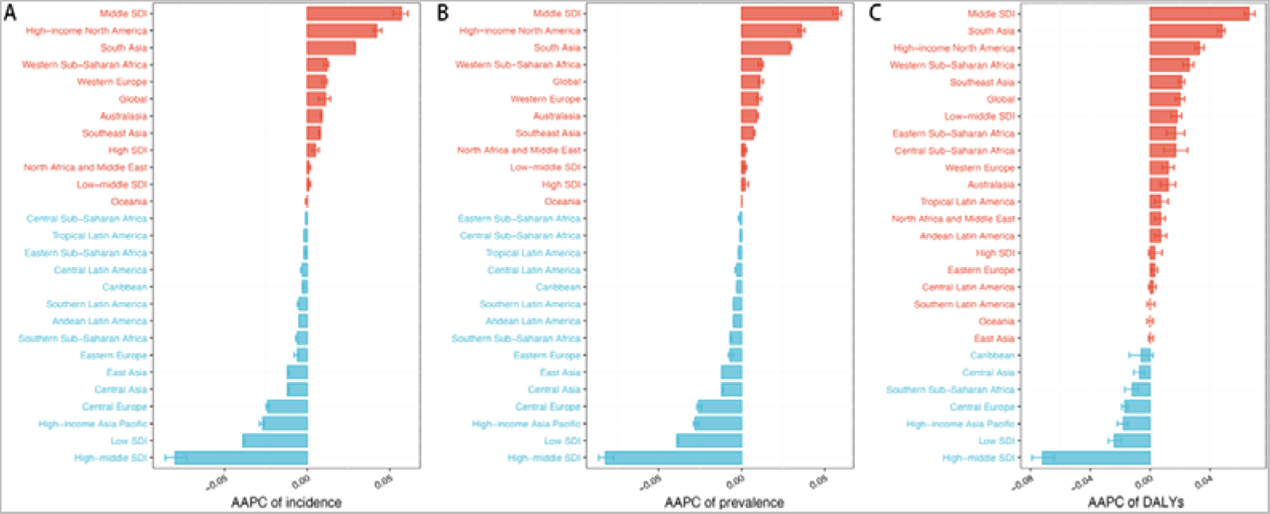
AAPC of the age-standardized incidence, age-standardized prevalence, age-standardized DALYs for urticaria from 1990 to 2019. Red represents values greater than 0 and blue represents values less than 0. (A) AAPC of incidence. (B)AAPC of prevalence. (C)AAPC of DALYs.

**Table 1.**
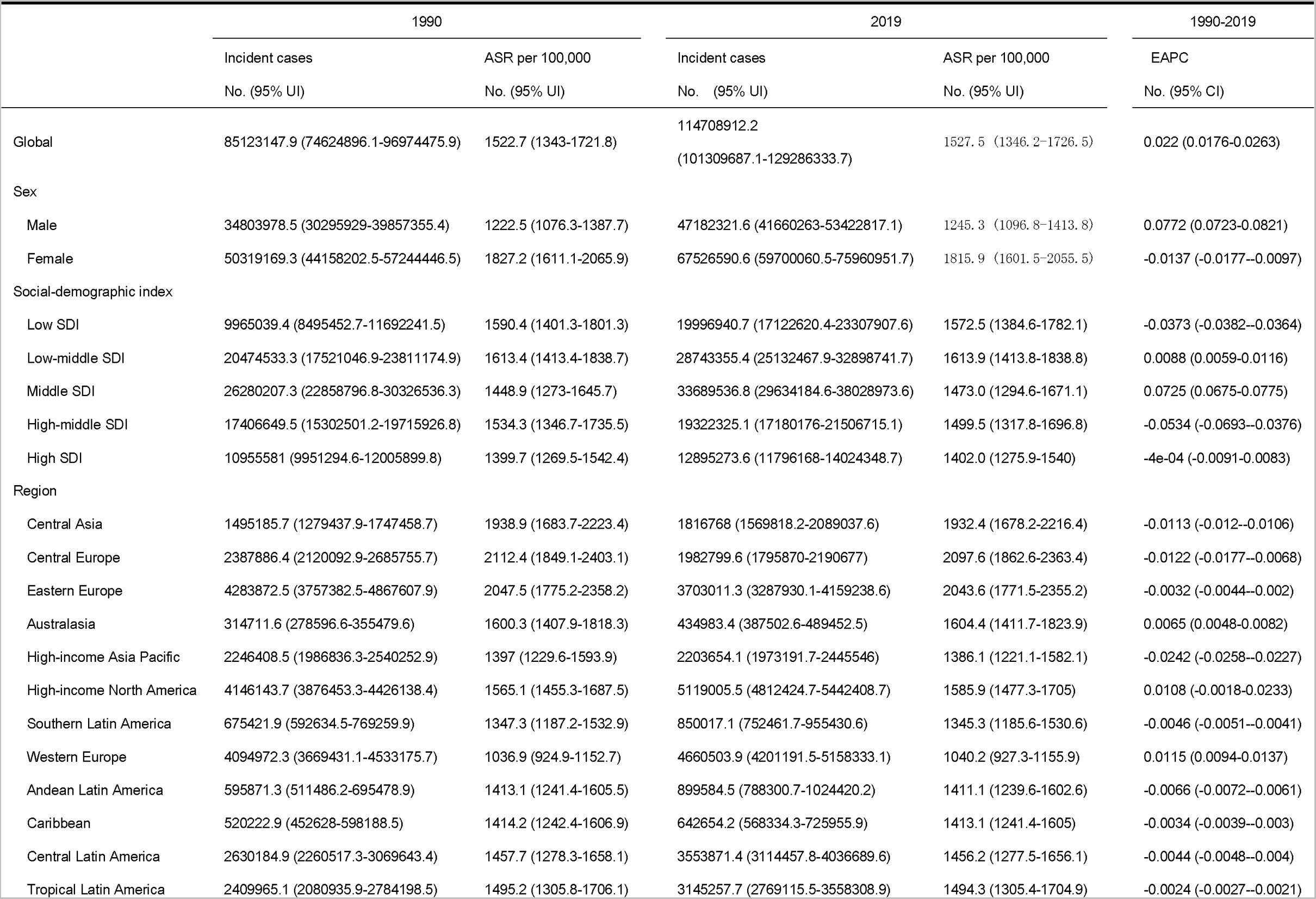
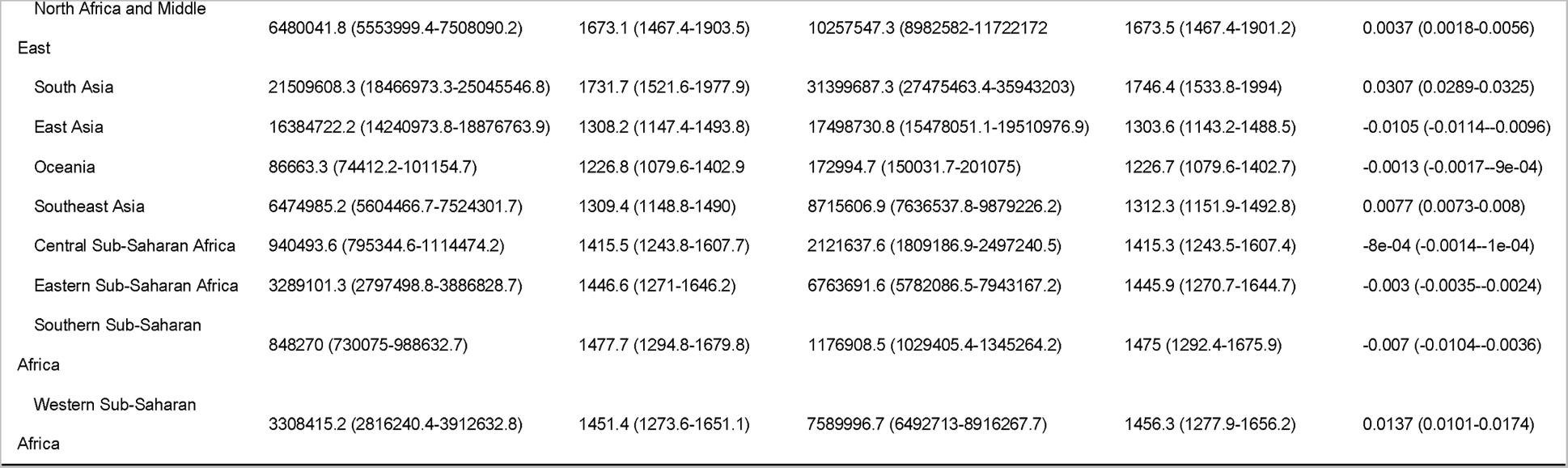
The incidence case and ASR per 100,000 of urticaria in 1990 and 2019, and the EAPC of incidence spanning 1990-2019.

Simultaneously, there were 65139886.6 (95% UI=57516618-73498729.7) prevalence cases globally in 2019, with an age-standardized prevalence rate (ASPR) equating to 865.5 (95% UI=761.8-980.6) per 100,000 population. The EAPC (0.0234 (0.0192-0.0276)) and AAPC of prevalence both exceeded 0, thereby showing an upward trend in the ASPR spanning from 1990 to 2019 (Table 2, Figure 1B).

**Table 2.**
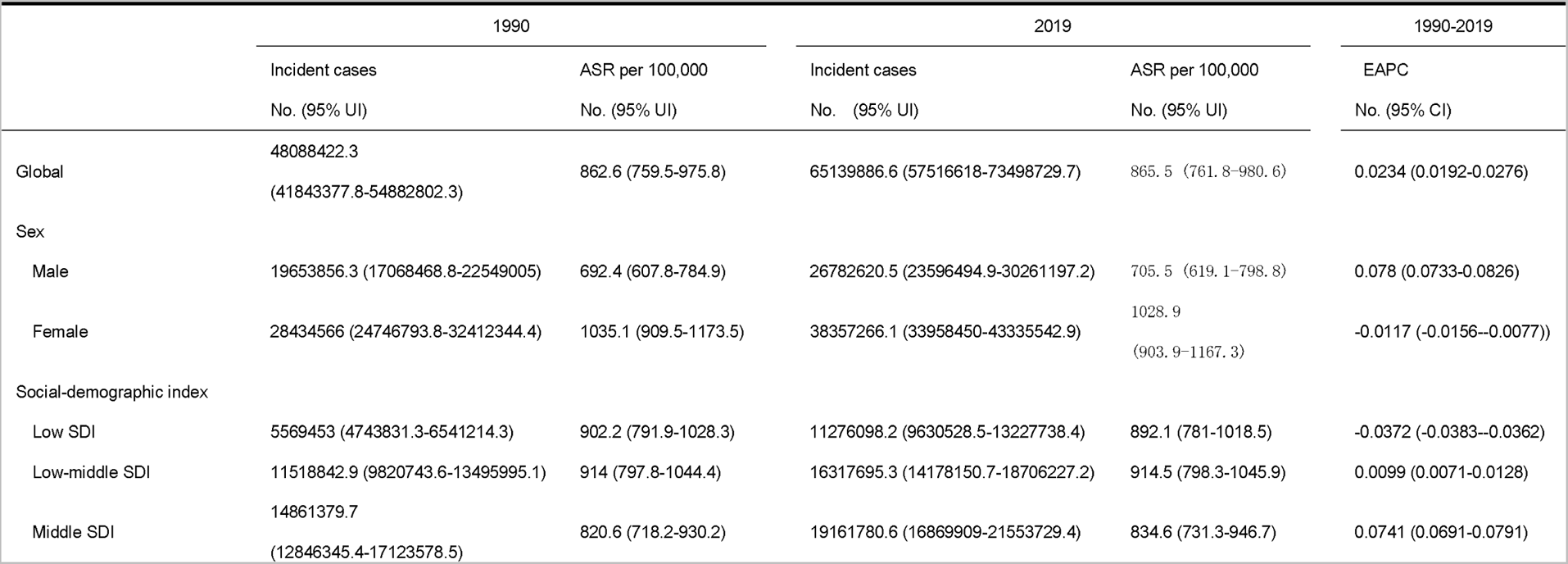
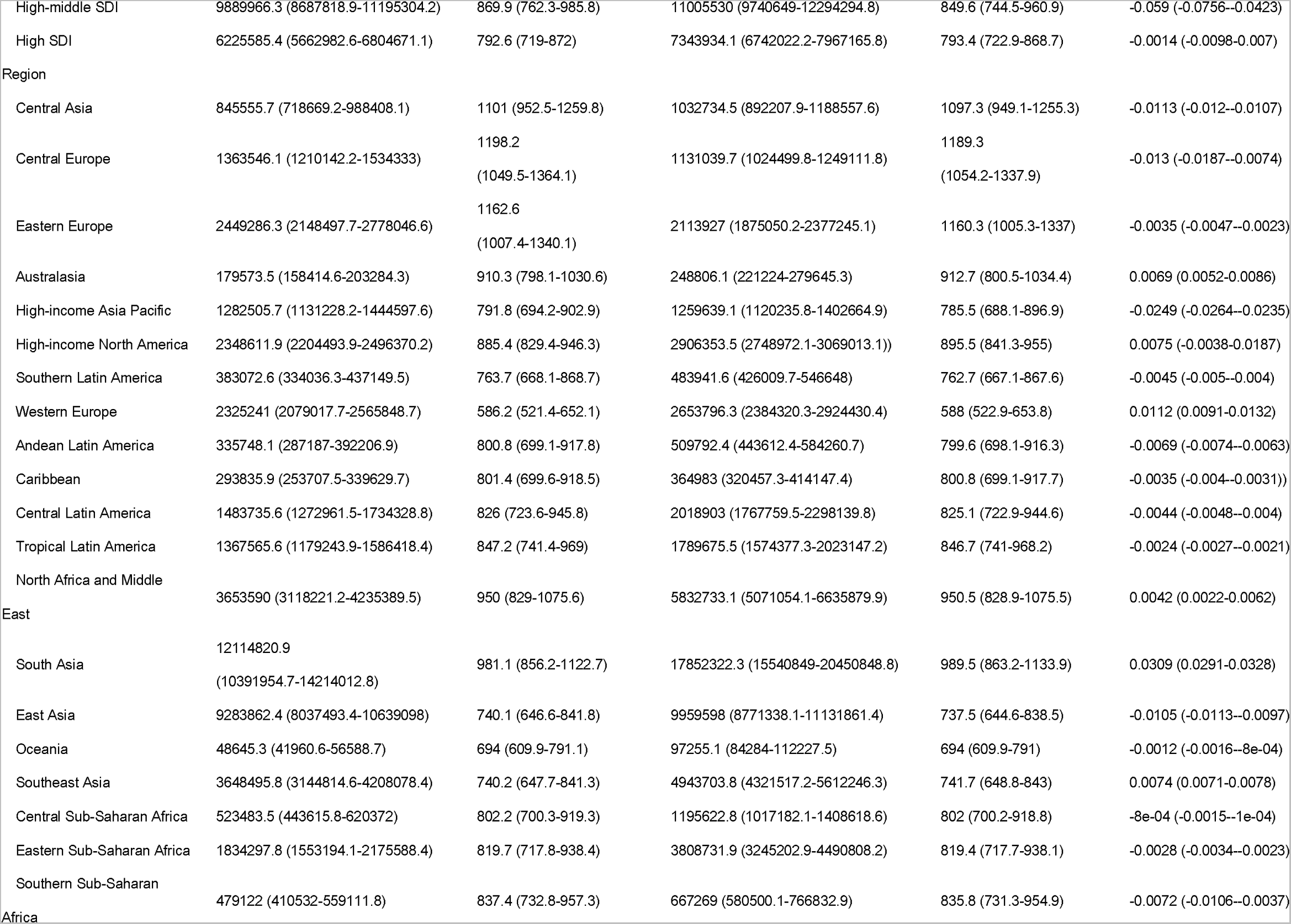
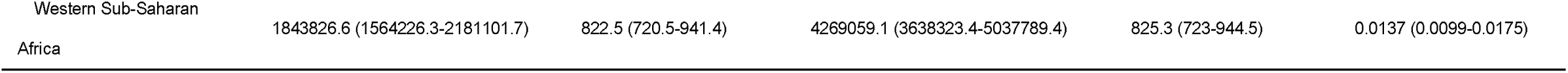
The prevalence case and ASR per 100,000 of urticaria in 1990 and 2019, and the EAPC of prevalence spanning 1990-2019.

Regarding DALYs, the global burden attributable to urticaria measured 3898838.6 (95% UI: 2554225.4-5584365.6), and age-standardized of DALYs measured 51.9 (95% UI: 34-75.1) per 100,000 population, and both EAPC (0.0338 (0.0287-0.0388)) and AAPC of DALYs were both greater than 0, thereby showing an upward trend from 1990 to 2019 (Table 3, Figure 1C).

**Table 3.** The DALY case and ASR per 100,000 of urticaria in 1990 and 2019, and the EAPC of DALY spanning 1990-2019.

|  | 1990 |  | 2019 |  | 1990-2019 |
| --- | --- | --- | --- | --- | --- |
|  | DALY cases | ASR per 100,000 | DALY cases | ASR per 100,000 | EAPC |
|  | No. (95% UI) | No. (95% UI) | No. (95% UI) | No. (95% UI) | No. (95% CI) |
| Global | 2889695.3 (1881485.9-4200997.5) | 51.6 (33.8-74.2) | 3898838.6 (2554225.4-5584365.6) | 51.9 (34-75.1) | 0.0338 (0.0287-0.0388) |
| Sex |  |  |  |  |  |
| Male | 1187604.9(776134.6-1720442.3) | 41.6(27.3-60.1) | 1613269.8(1051942.8-2320511.4) | 42.5(27.8-61.7) | 0.0878(0.0823-0.0932) |
| Female | 1702090.3(1112433.9-2468057.8) | 61.7(40.5-88.3) | 2285568.8(1499168.1-3272442.3) | 61.5(40.4-88.8) | -0.0013(-0.0061-0.0036) |
| Social-demographic index |  |  |  |  |  |
| Low SDI | 334010.1 (218227.5-489455.3) | 53.5 (35.1-77.4) | 678840.9 (443143.6-992715.1) | 53.2 (34.9-76.6) | -0.0173 (-0.0196--0.0149) |
| Low-middle SDI | 692177.7 (450787.7-1008459.3) | 54.4 (35.6-78.5) | 979311.1 (639278.3-1420447.6) | 54.7 (35.8-79.1) | 0.0281 (0.0243-0.032) |
| Middle SDI | 897567 (584515.9-1299659.9) | 49.2 (32.1-70.7) | 1149480.9 (752418-1643055.5) | 50.2 (32.8-72.2) | 0.0836 (0.078-0.0892) |
| High-middle SDI | 594152.8 (387212.6-856327.3) | 52.2 (34.1-75.2) | 656089.3 (432160.1-936970.6) | 51.2 (33.5-73.7) | -0.047 (-0.0638--0.0301) |
| High SDI | 370391.8 (244053.2-522773.6) | 47.5 (31.2-67.4) | 433027.6 (285343.5-609777.2) | 47.5 (31.3-67.6) | -4e-04 (-0.0103-0.0095) |
| Region |  |  |  |  |  |
| Central Asia | 51231.8 (33493.6-74614.6) | 66.3 (43.6-95.8) | 62416.7 (40782.9-90250) | 66.2 (43.3-96) | -0.005 (-0.0063--0.0036) |
| Central Europe | 81794.8 (53933.2-118171.5) | 72.3 (47.5-104.2) | 67169.3 (44596.1-95460.2) | 71.9 (47.2-103.4) | -0.0034 (-0.0084-0.0017) |
| Eastern Europe | 146607.4 (95797.1-211523.6) | 70 (45.7-102.1) | 125843.9 (82383.2-180930.6) | 70.1 (46-102.1) | 0.0068 (0.005-0.0085) |
| Australasia | 10667.4 (6964.2-15115.8) | 54.3 (35.7-77.5) | 14694.8 (9608.9-21046.6) | 54.5 (35.3-78.2) | 0.011 (0.0076-0.0144) |
| High-income Asia Pacific | 76764.3 (50531.2-110521.8) | 47.7 (31.3-69) | 74570 (49254.8-105635.9) | 47.4 (31.1-68.4) | -0.0149 (-0.0172--0.0126) |
| High-income North America | 139663.5 (92687.9-197132.9) | 53 (35.1-74.9) | 171109 (114326.9-239859.8) | 53.5 (35.5-75.6) | 0.0018 (-0.0116-0.0153) |
| Southern Latin America | 23038.1 (15073.3-33464) | 45.9 (29.9-66.5) | 28928.2 (18987.6-41298.2) | 45.8 (30-66.2) | 1e-04 (-0.0024-0.0026) |
| Western Europe | 136808.9 (90836.4-191363) | 34.8 (22.9-49.1) | 155143.4 (103509.7-217649.1) | 34.9 (23-49.4) | 0.0161 (0.0135-0.0187) |
| Andean Latin America | 20290.1 (13291.1-29655.9) | 48 (31.5-69.2) | 30723.5 (19985.1-44502.2) | 48.1 (31.4-69.8) | 0.0078 (0.0055-0.0101) |
| Caribbean | 17706.5 (11547.5-25817.9) | 48.1 (31.5-69.8) | 21826.7 (14363.9-31358.7) | 48 (31.5-69.1) | -0.0069 (-0.0084--0.0054) |
| Central Latin America | 89957.4 (58494.7-131864.7) | 49.6 (32.3-72) | 121438.7 (79078.5-175939.7) | 49.7 (32.4-72.3) | 0 (-0.0013-0.0013) |
| Tropical Latin America | 82324.2 (53547.9-119971.2) | 50.6 (33.2-73.3) | 106852.1 (69841.2-154007.6) | 50.8 (33.1-73.5) | 0.0073 (0.0052-0.0093) |
| North Africa and Middle East | 220985.6 (144152.2-323560.9) | 56.9 (37.1-82) | 350946.2 (229607.5-508231.2) | 57 (37.4-82.8) | 0.0094 (0.0077-0.0111) |
| South Asia | 726277 (473731.7-1055685.6) | 58.3 (38.3-83.8) | 1068940.9 (696805.2-1546075.7) | 59.1 (38.6-85.3) | 0.0518 (0.0497-0.054) |
| East Asia | 561250 (364288-810674.3) | 44.6 (29.2-64.7) | 595886.5 (392144.5-845623.1) | 44.6 (29.1-64.8) | 0.0022 (6e-04-0.0039) |
| Oceania | 2934.5 (1910.8-4278) | 41.5 (27.2-59.3) | 5853.3 (3843.8-8629.9) | 41.4 (27.1-59.8) | 2e-04 (-0.0018-0.0023) |
| Southeast Asia | 220182.8 (145247.1-315169.9) | 44.3 (29.3-63.4) | 297242.9 (196270.5-423283.2) | 44.6 (29.4-63.7) | 0.0254 (0.0236-0.0271) |
| Central Sub-Saharan Africa | 31361.3 (20644.9-46005.1) | 47.5 (31.4-68.7) | 71958.9 (46757.3-105267.1) | 47.8 (31.5-68.9) | 0.0219 (0.0178-0.0261) |
| Eastern Sub-Saharan Africa | 110306.9 (71728.9-162984.1) | 48.8 (31.8-70.8) | 230139.9 (150650.3-338951) | 49 (32.1-71.1) | 0.0238 (0.0208-0.0269) |
| Southern Sub-Saharan Africa | 28938.1 (18881.3-42307.8) | 50.1 (32.8-72.7) | 40035 (25993.7-58297.4) | 50 (32.6-72.5) | -0.0101 (-0.0115--0.0088) |
| Western Sub-Saharan Africa | 110604.6 (72061.7-162297.9) | 48.9 (32-70.8) | 257118.5 (168026.7-376269.7) | 49.3 (32.3-71.5) | 0.031 (0.0247-0.0373) |

### 3.2 Regional urticaria burden

According to the GBD database, the world is divided into 21 GBD Regions. With respect to the ASIR, the highest rate in 2019 appeared in Central Europe (2097.6 (1862.6-2363.4)), Eastern Europe (2043.6 (1771.5-2355.2)), Central Asia (1932.4 (1678.2-2216.4)), with the lowest rate in East Asia (1303.6 (1143.2-1488.5)), Oceania (1226.7 (1079.6-1402.7)), Western Europe (1040.2 (927.3-1155.9)). Concerning the EAPC of incidence, the strongest upward trend was identified in South Asia (0.0307 (0.0289-0.0325)), while the strongest downward trend region is High-income Asia Pacific (−0.0242 (−0.0258--0.0227)) (Table 1).

In the ASPR part, the apogee was observed in Central Europe (1189.3 (1054.2-1337.9)), Eastern Europe (1160.3 (1005.3-1337)) and Central Asia (1097.3 (949.1-1255.3)), with the lowest rate in East Asia (737.5 (644.6-838.5)), Oceania (694 (609.9-791)) and Western Europe (588 (522.9-653.8)). Concerning the EAPC of prevalence, the strongest upward trend was identified in South Asia (0.0309 (0.0291-0.0328)), while the strongest downward trend region is High-income Asia Pacific (−0.0249 (−0.0264--0.0235)) (Table 2).

In the age-standardized of DALYs part, the apogee was observed in Central Europe (71.9 (47.2-103.4)), Eastern Europe (70.1 (46-102.1)) and Central Asia (66.2 (43.3-96)), with the lowest rate in Western Europe (34.9 (23-49.4)), Oceania (41.4 (27.1-59.8)), and East Asia (44.6 (29.1-64.8)). Concerning the EAPC of DALYs, the strongest upward trend region was identified in South Asia (0.0518 (0.0497-0.054)), while the strongest downward trend region is High-income Asia Pacific (−0.0149 (−0.0172--0.0126)) (Table 3).

For the regional AAPC of the incidence, High-income North America exhibited the biggest upward trend, succeeded by South Asia, which also manifested an increasing trend. Conversely, Central Europe and High-income Asia Pacific exhibited the most significant downward trends. Similarly, the AAPC of the prevalence showed the biggest increasing trend in High-income North America and succeeded by South Asia, while Central Europe and High-income Asia Pacific exhibited the biggest decreasing trend. For the regional AAPC of DALYs, South Asia exhibited the biggest increasing trend and succeeded by High-income North America, while Central Europe and High-income Asia Pacific exhibited the biggest decreasing trend (Figure 1A-C).

### 3.3 National urticaria burden

At the national scale, the data for 2019 exhibit Nepal got the highest ASIR(2665.475289 UI /10^5), ASPR(1534.557178 UI /10^5), and the age-standardized of DALYs(92.01297809 UI /10^5) at the same time. Conversely, Portugal exhibited the lowest figures in these key indicators. (Figure 2A, Figure 3A, Figure 4A). The EAPC of the incidence has the biggest upward trend in San Marino, Andorra, Nepal, Bangladesh, Greenland, and the biggest decreasing trend in Qatar, Equatorial Guinea, Democratic People’s Republic of Korea, Malta, Cabo Verde (Figure 3B). This sequence similarly applies to the EAPC of prevalence and DALYs (Figure 2B, Figure 4B).

**Figure 2.**
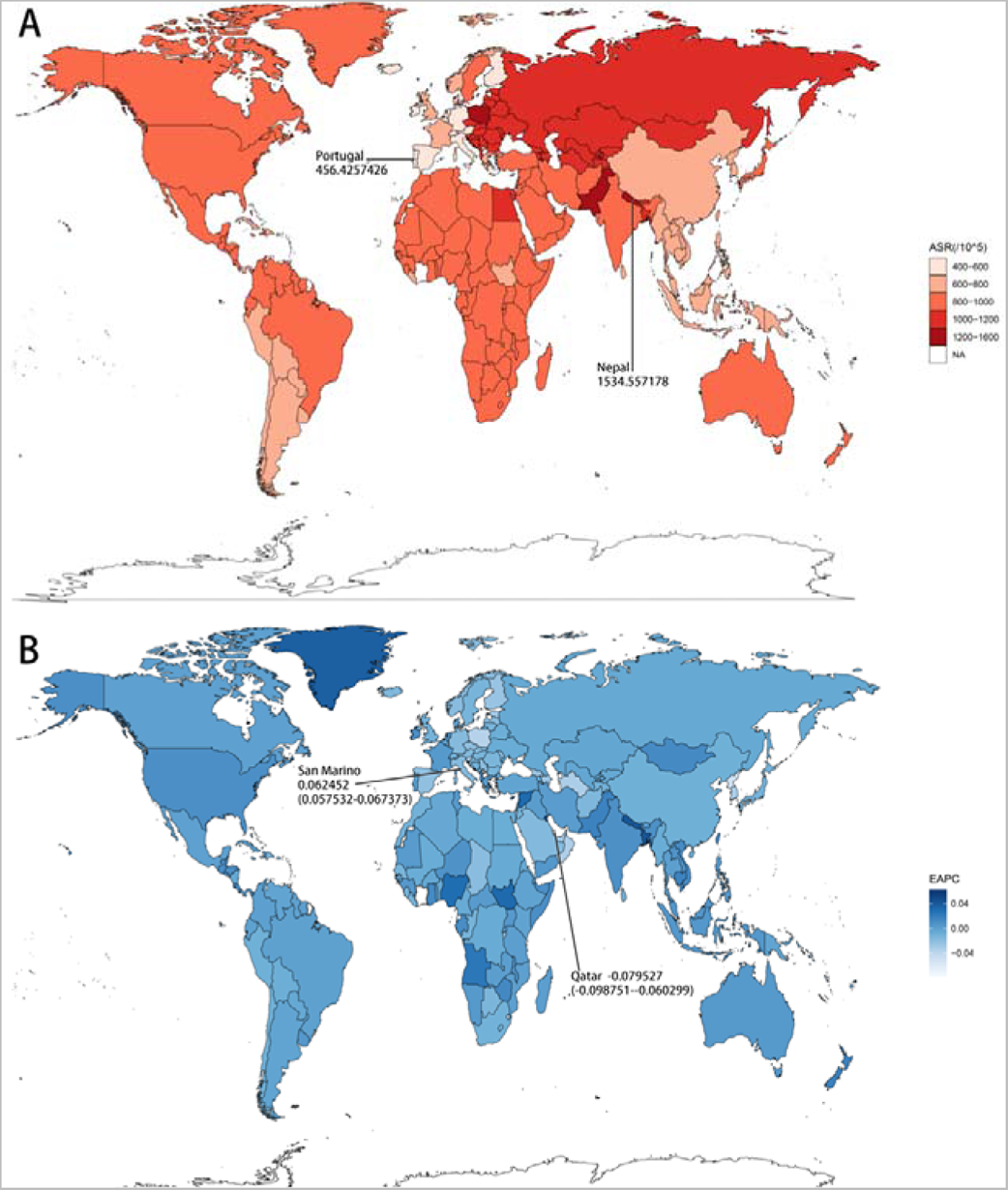
The urticaria global ASR (per 10^5) prevalence map and EAPC of prevalence map in 2019 by countries and territories. (A) ASR (per 10^5) DALY map. (B) EAPC of DALY map.

**Figure 3.**
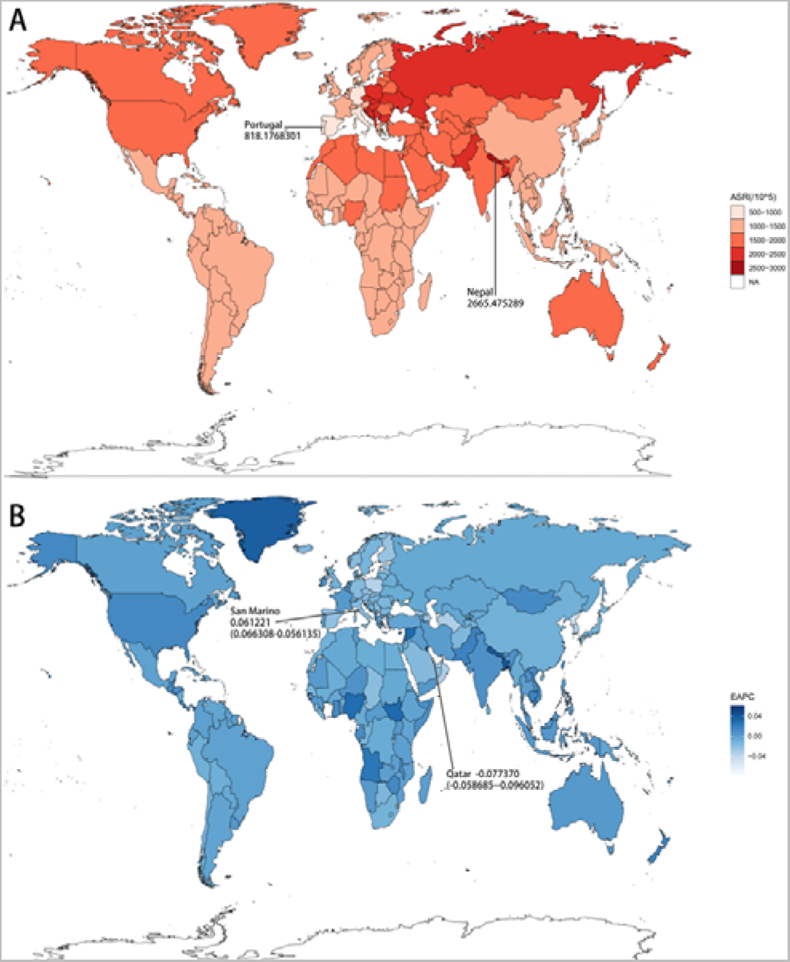
The urticaria global ASR (per 10^5) incidence map and EAPC of incidence map in 2019 by countries and territories. (A) ASR (per 10^5) DALY map. (B) EAPC of DALY map.

**Figure 4.**
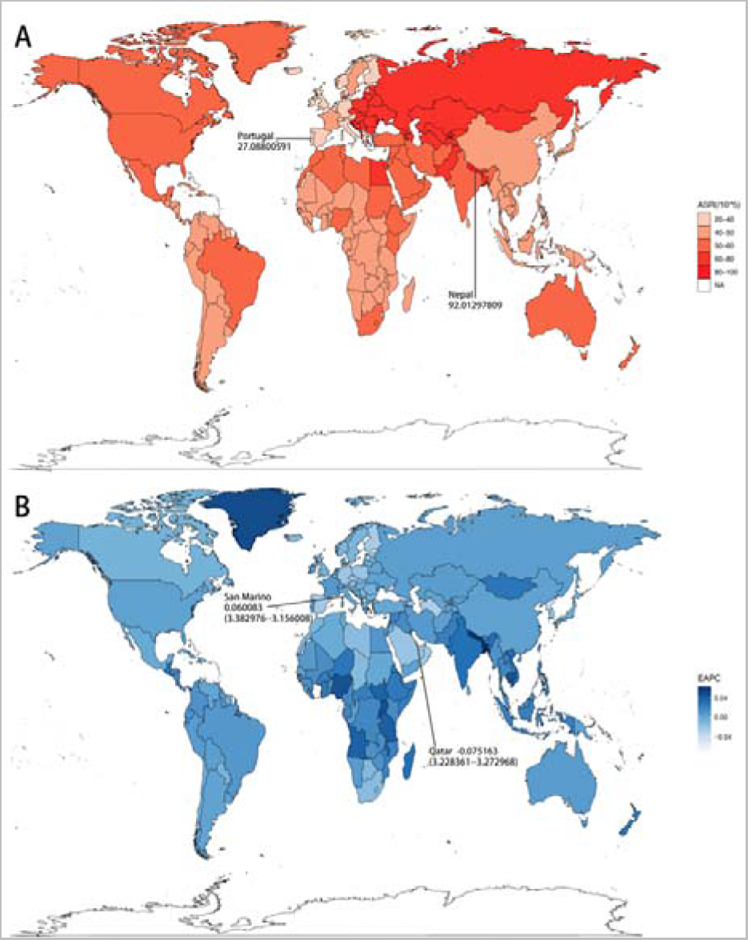
The urticaria global ASR (per 10^5) DALY map and EAPC of DALY map in 2019 by countries and territories. (A) ASR (per 10^5) DALY map. (B) EAPC of DALY map.

### 3.4 Age and sex

The data indicate a decline in urticaria incidence and prevalence concomitant with age growth, especially within the 1-14 years old. Thereafter, the decline tended to weaken gradually, yet a slight resurgence is observed between 50 and 69 years old. Gender-specific analysis shows that EAPCs for incidence, prevalence, and DALYs for females are negative values, meaning a declining trend, while for males are positive values, indicating an upward trend (Figure 5–7, Table 1–3).

**Figure 5.**
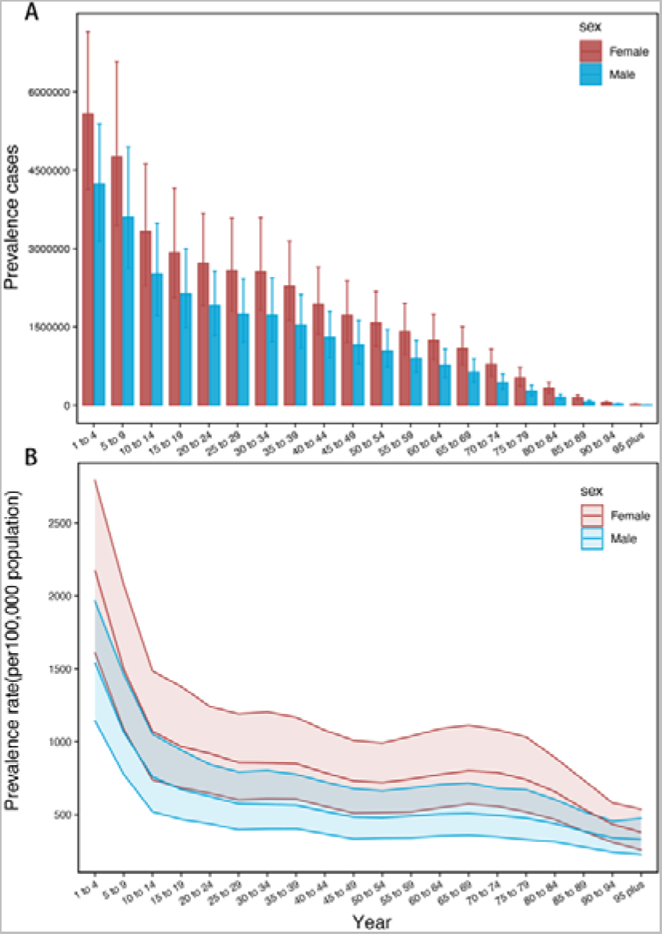
The prevalence cases and prevalence rates(per100,000 population) of urticaria among age and gender in 2019. (A) Prevalence cases. (B) Prevalence rates.

**Figure 6.**
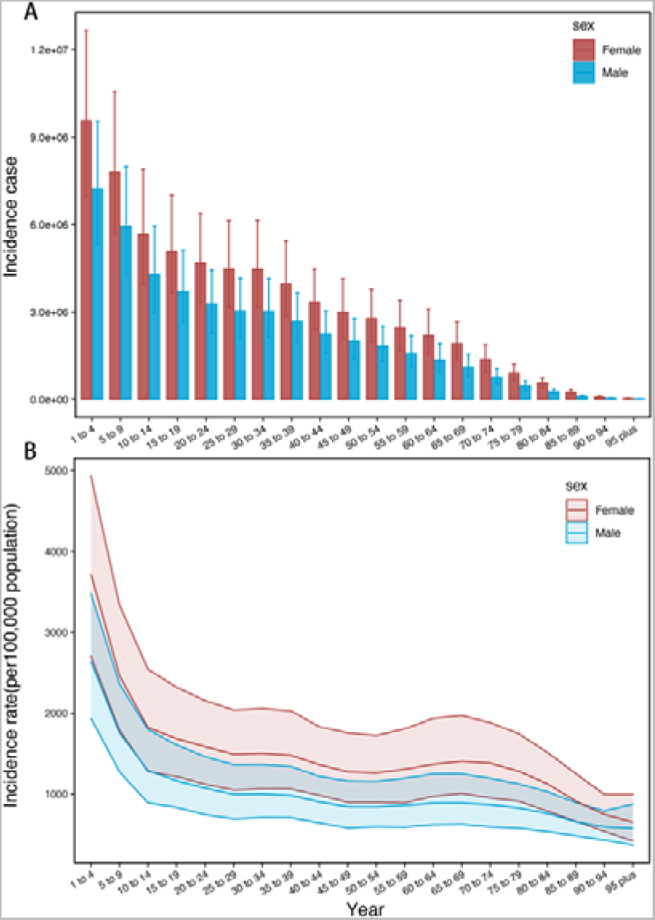
The incidence cases and incidence rates(per100,000 population) of urticaria among age and gender in 2019. (A) Incidence cases. (B) Incidence rates.

**Figure 7.**
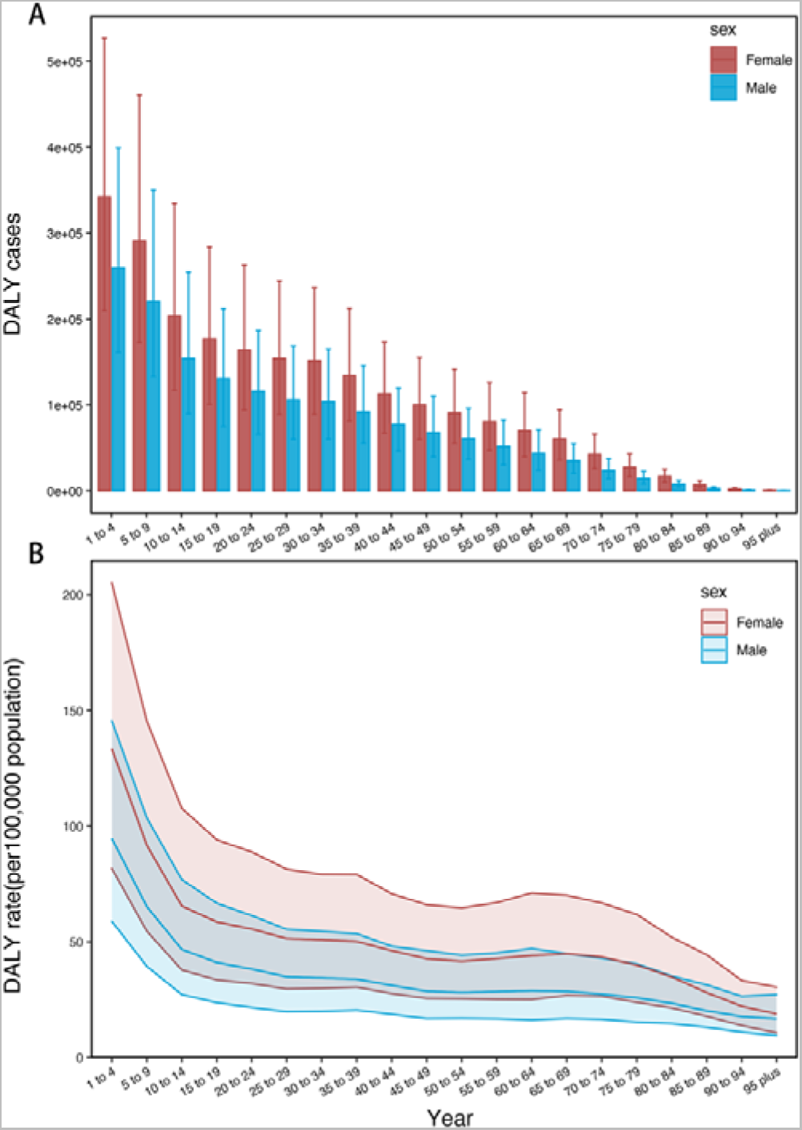
The DALY cases and DALY rates(per100,000 population) of urticaria among age and gender in 2019. (A) DALY cases. (B) DALY rates.

### 3.5 Socio-demographic Index (SDI)

In the relationship between incidence and regional SDI, ASIR exhibits a slightly positive correlation with regional SDI (R=0.1, p <0.01). Specifically, regions such as Central Europe, Eastern Europe, Central Asia, and South Asia deviate above the fitted curve, indicating that their actual values surpass the expected. Conversely, regions like Western Europe, Oceania, East Asia, Southeast Asia, Southern Latin America fall below the fitted curves, indicating that their actual values are less than projected (Figure 9A). For ASIR correlated with country-level SDI, there was no obvious trend (p >0.05) (Figure 9B). In the fitted curve between incidence EAPC and SDI, there was a negative correlation (R=-0.2, p<0.01) (Figure 8A).

**Figure 8.**
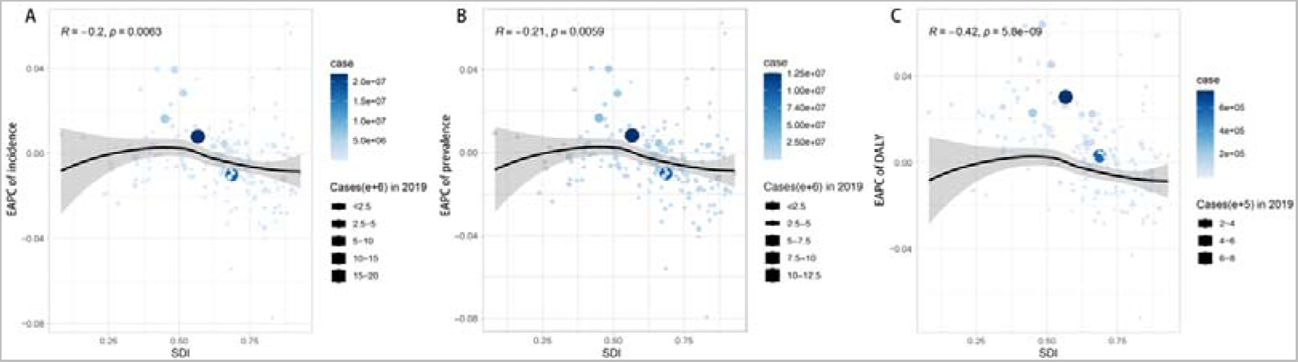
The correlation between urticaria EAPC and SDI levels in 2019. (A)EAPC of ASIR and SDI correlation. (B) EAPC of ASPR and SDI correlation. (C) EAPC of DALY and SDI correlation.

**Figure 9.**
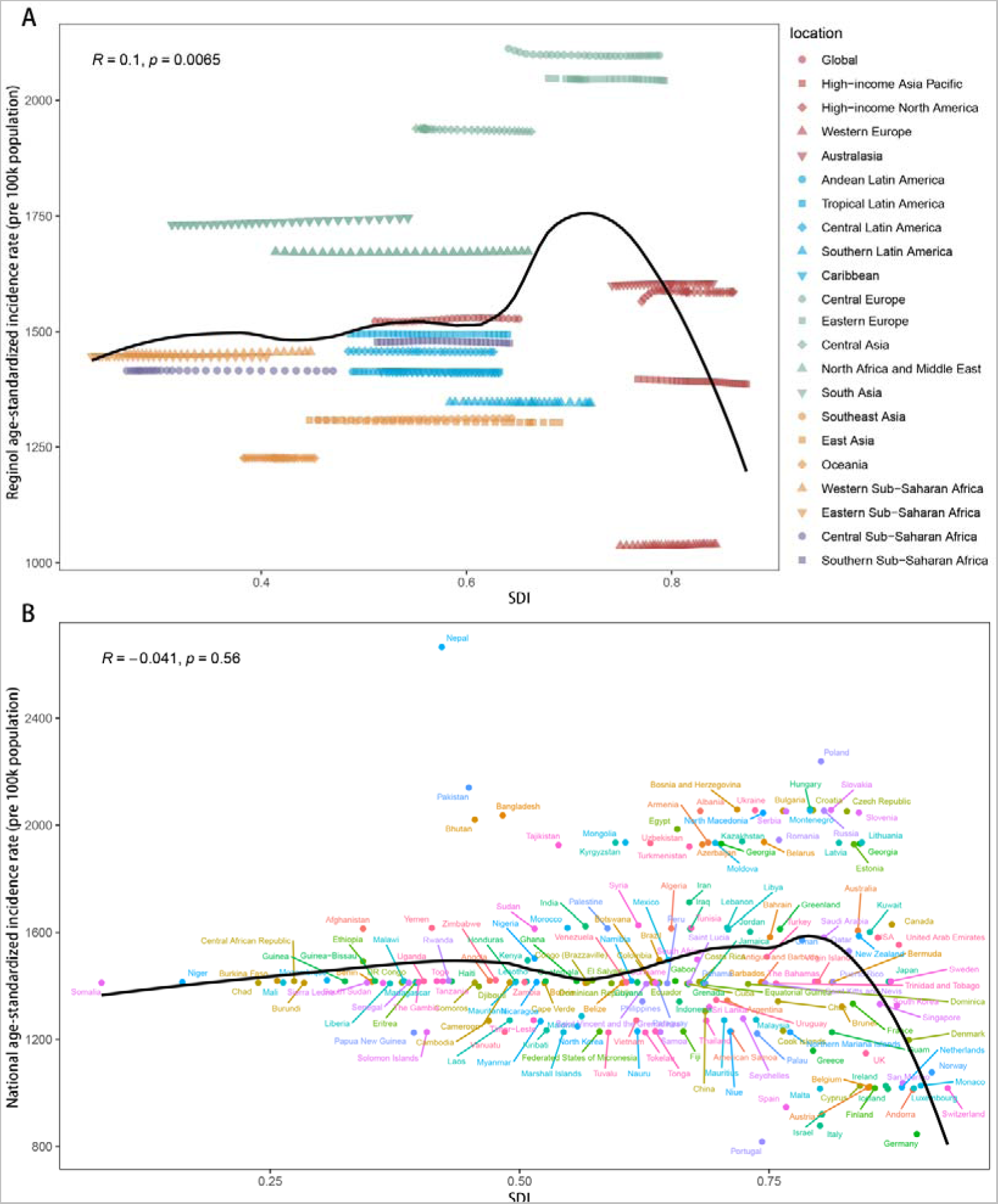
The correlation between urticaria ASIR and SDI levels in 2019. (A)Regional ASIR and SDI correlation. (B) National ASIR and SDI correlation.

In the association between prevalence and regional SDI, ASPR is slightly positively correlated with regional SDI (R=0.1, p <0.01). Similar to the incidence, Central Europe, Eastern Europe, Central Asia, and South Asia exceeded the fitted curve, while Western Europe, Oceania, East Asia, Southeast Asia, Southern Latin America fell below the fitted curves (Figure 10A). For ASPR and country SDI, there was no obvious trend (p >0.05) (Figure 10A). The fitted curve between the prevalence EAPC and SDI showed a negative correlation (R=-0.21, p<0.01) (Figure 8B).

**Figure 10.**
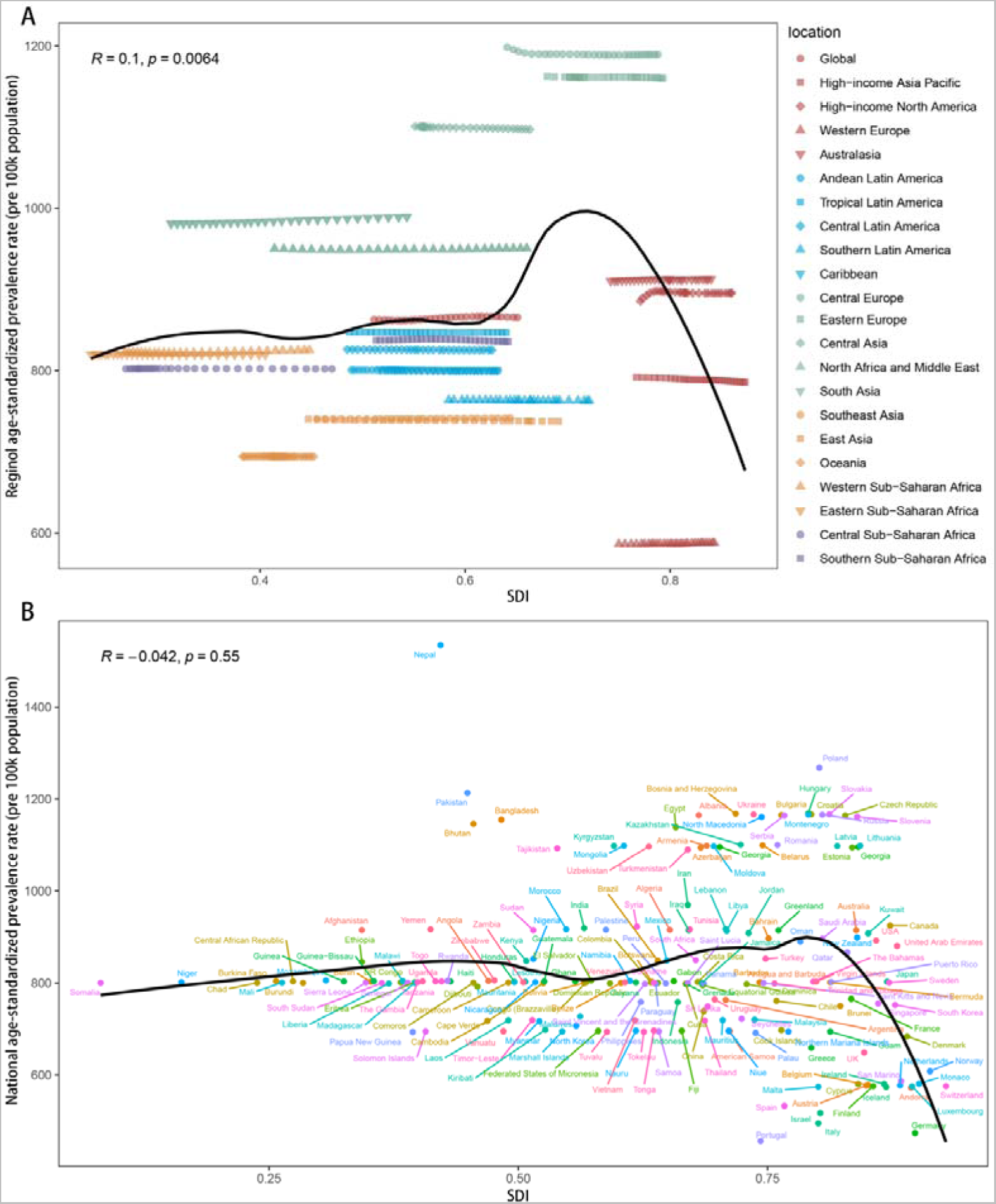
The correlation between urticaria ASPR and SDI levels in 2019. (A)Regional ASPR and SDI correlation. (B) National ASPR and SDI correlation.

Concerning the relationship between DALYs and regional SDI, age-standardized DALYs is slightly positively correlated with regional SDI (R=0.12, p <0.01). Regions such as Central Europe, Eastern Europe, and Central Asia exceeded the fitted curve, while Western Europe, Oceania, East Asia, Southeast Asia, Southern Latin America fell below the fitted curves (Figure 11A). For the age-standardized DALYs and country SDI, there was no obvious trend (p >0.05) (Figure 11A). In the fitted curve between DALYs EAPC and SDI, they are significantly negatively correlated (R=-0.42, p<0.01) (Figure 8C).

**Figure 11.**
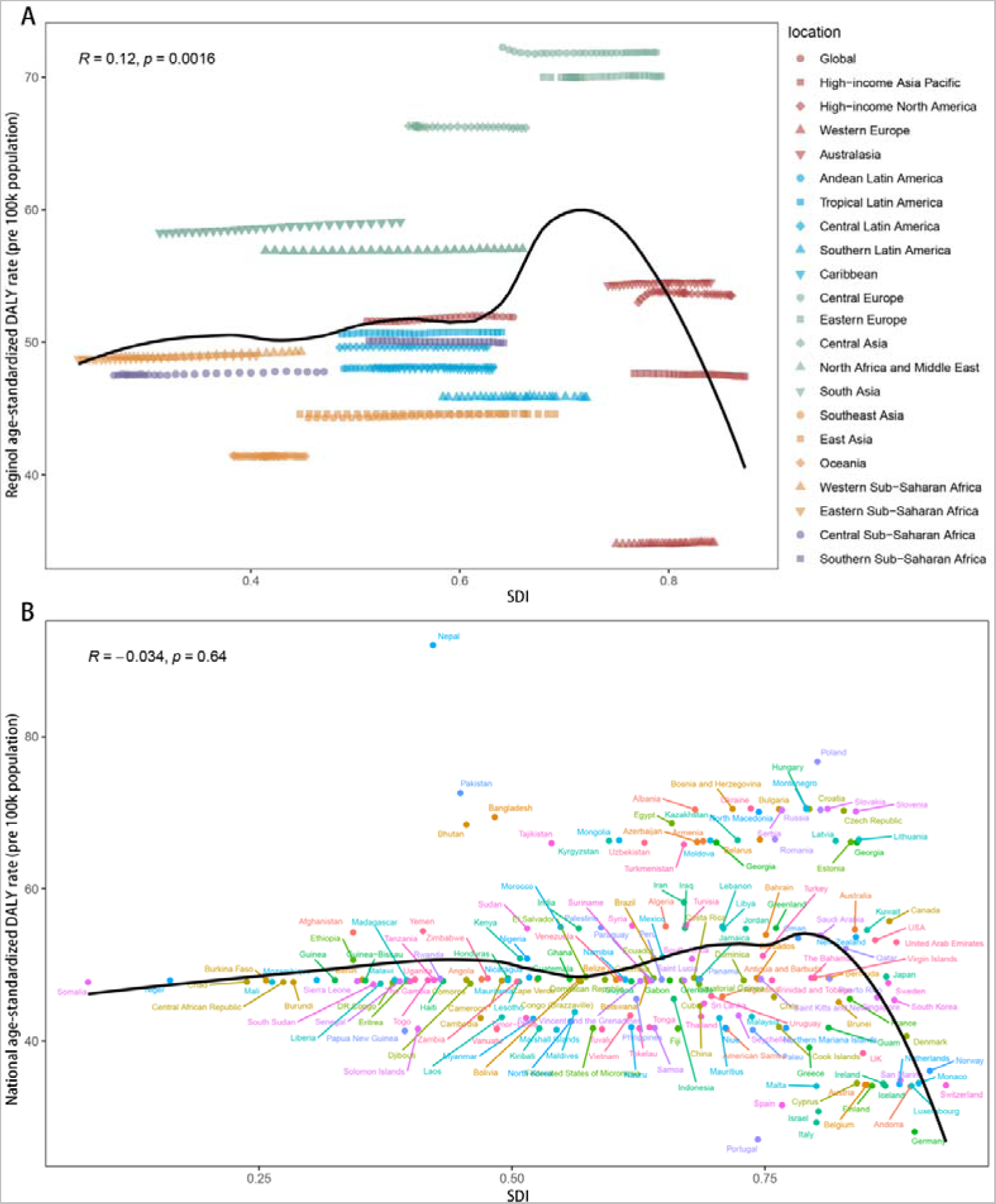
The correlation between urticaria age-standardized DALY and SDI levels in 2019. (A)Regional age-standardized DALY and SDI correlation. (B) National age-standardized DALY and SDI correlation.

Simultaneously, SDI was stratified into 5 different levels to analysis: low SDI, low-middle SDI, middle SDI, high-middle SDI, and high SDI. The incidence, prevalence, and DALY cases in the low-middle and middle SDI were significantly higher than those in other SDI strata. The main cases across all SDI categories were concentrated in the 15-49 age bracket, albeit the prevalence of cases under age 14 was higher in low, low-middle, and middle SDI compared to high-middle and high SDI categories (Figure 12-14).

**Figure 12.**
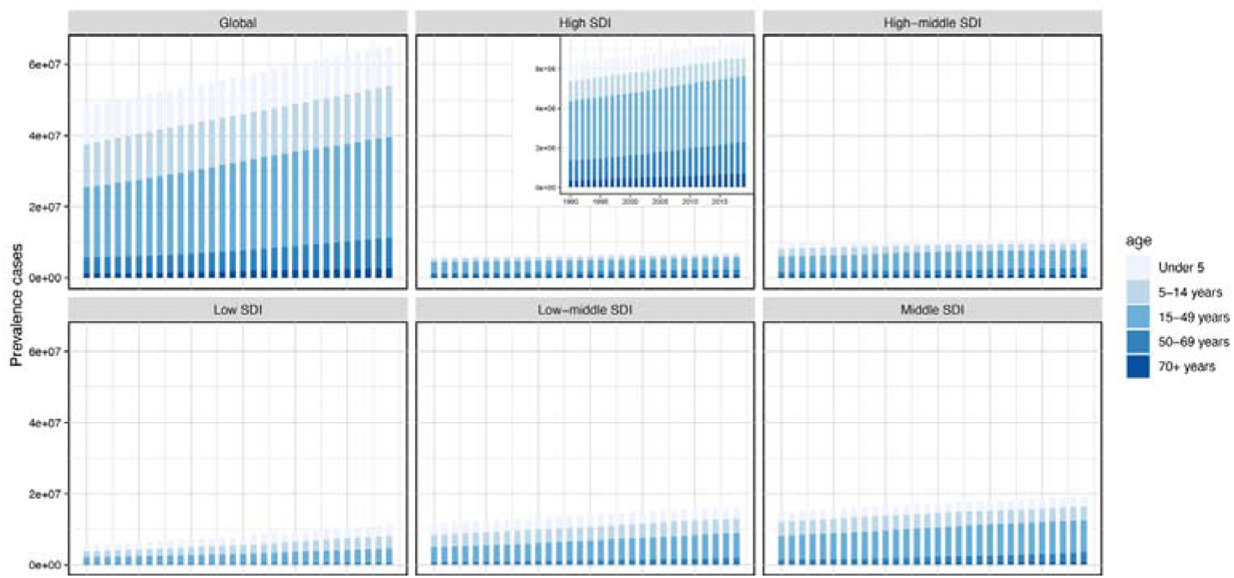
The urticaria prevalence cases of 5 age group in Global and 5SDI level from 1990 to 2019.

**Figure 13.**
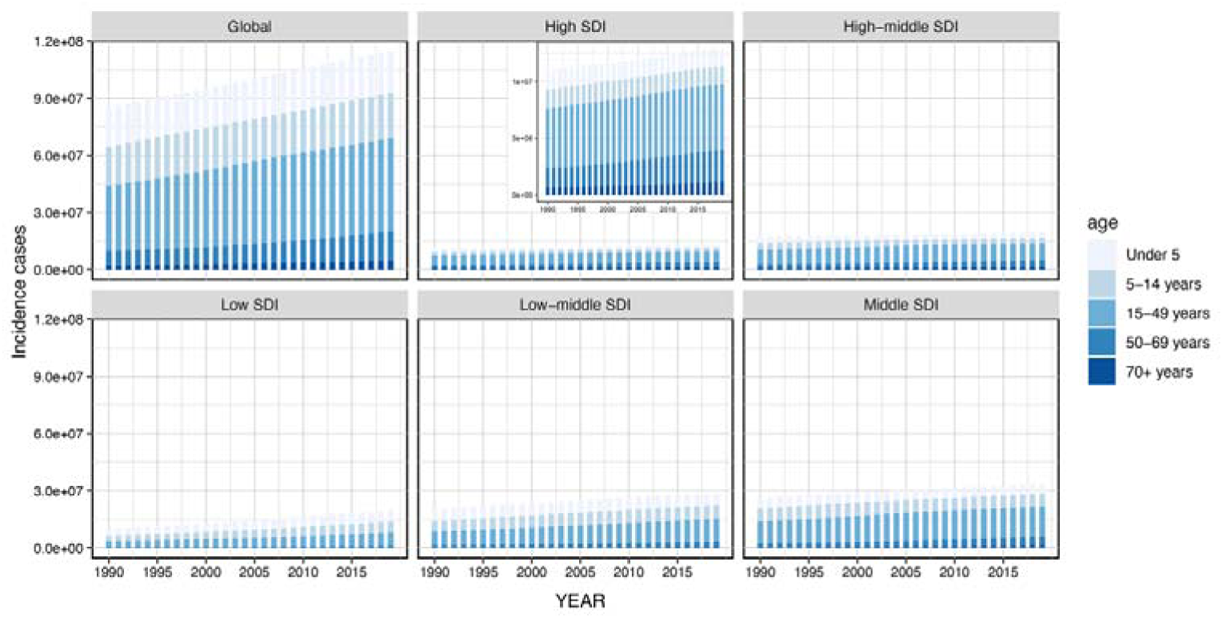
The urticaria incidence cases of 5 age group in Global and 5SDI level from 1990 to 2019.

**Figure 14.**
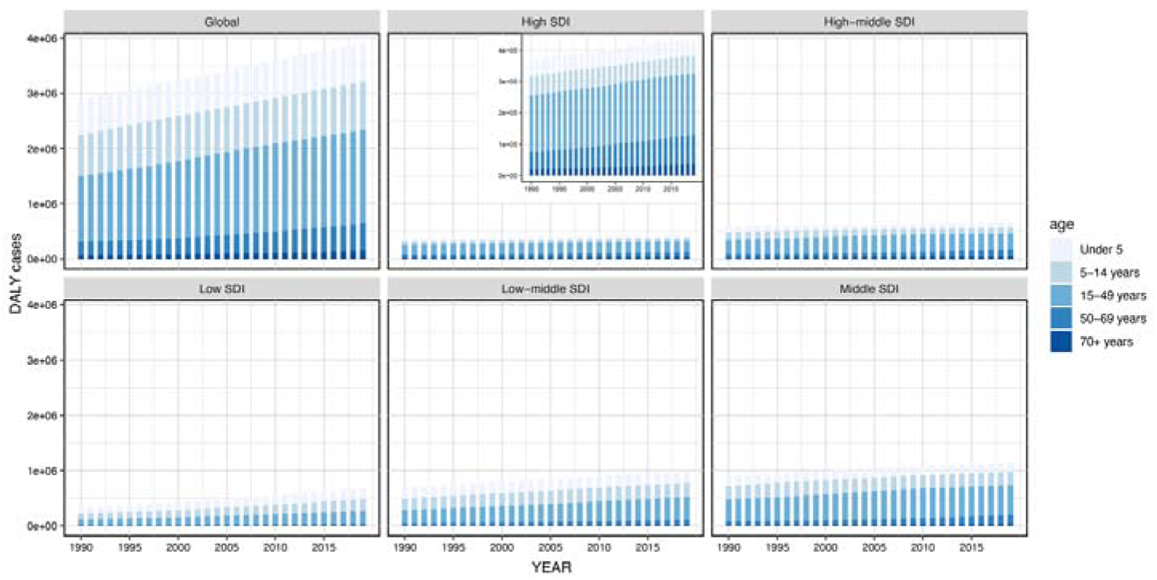
The urticaria DALY cases of 5 age group in Global and 5SDI level from 1990 to 2019.

In the joinpoint analysis spanning from 1990 to 2019, both ASIR and ASPR in low SDI and high-middle SDI categories displayed a continuous decline, whereas a consistent increase was observed in middle SDI. By contrast, the remaining SDI categories showed fluctuating trends (Figure 15, 16). In terms of DALYs, a downward trend was observed in low and high-middle SDI, while low-middle and middle exhibited an upward trend; high SDI showed a fluctuating trend (Figure 17). AAPC, derived from joinpoint analysis, showed that middle SDI got the highest among all AAPC values for incidence, prevalence, and DALYs, while the lowest was noted in high-middle and low SDI (Figure 1A-C).

**Figure 15.**
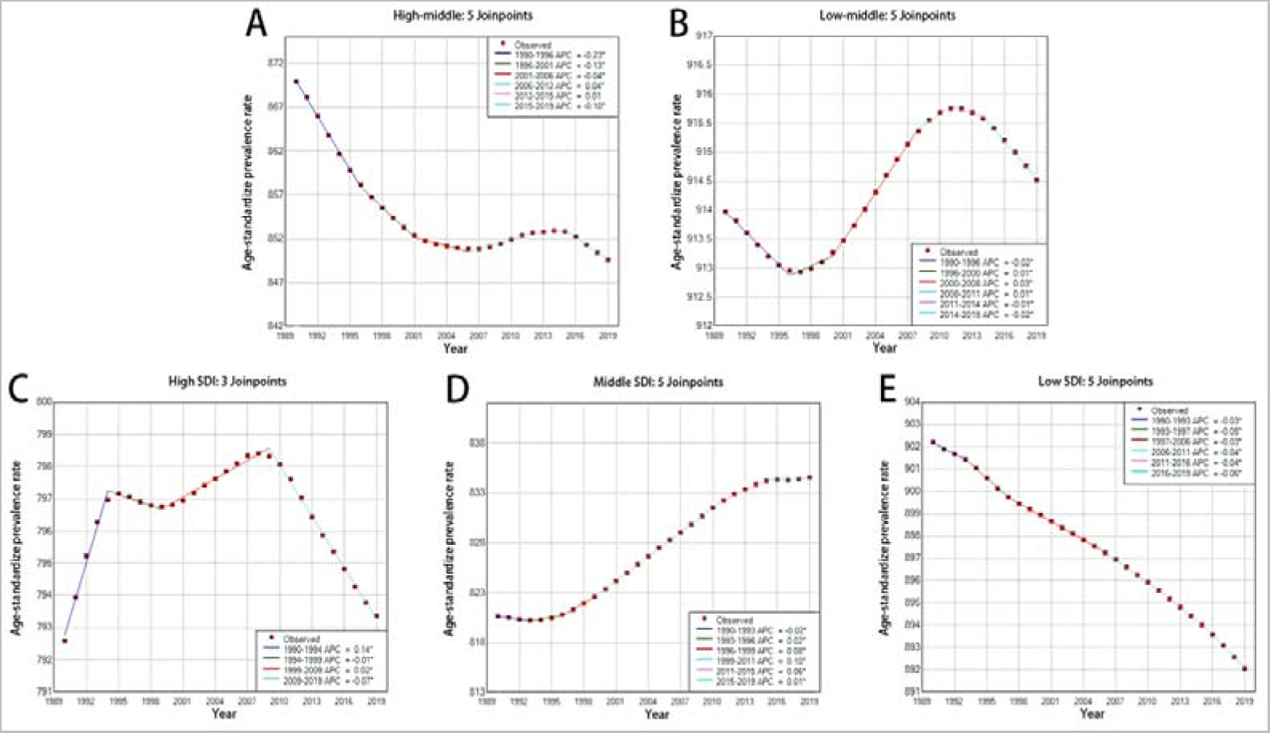
Temporal trends of ASPR in 5SDI levels by Jointpoint regression model. (A) High-middle SDI. (B) Low-middle SDI. (C) High SDI. (D) Middle SDI. (E) Low SDI.

**Figure 16.**
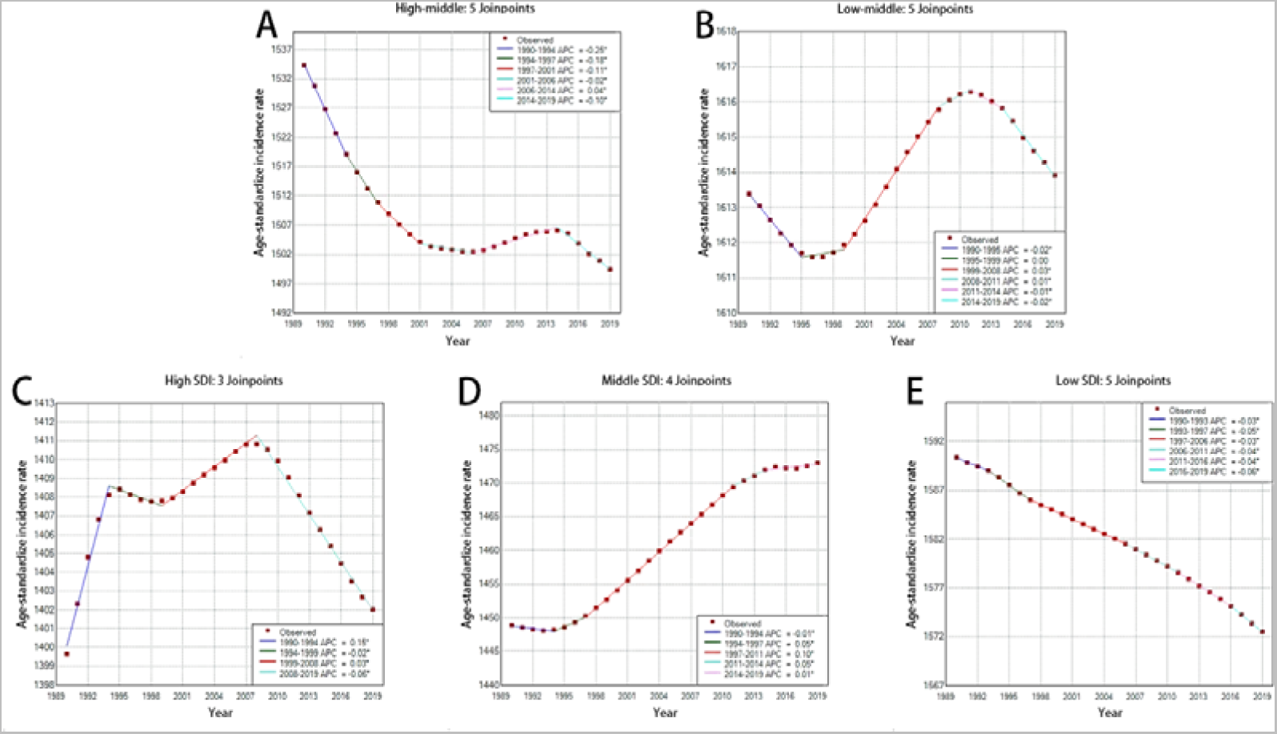
Temporal trends of ASIR in 5SDI levels by Jointpoint regression model. (A) High-middle SDI. (B) Low-middle SDI. (C) High SDI. (D) Middle SDI. (E) Low SDI.

**Figure 17.**
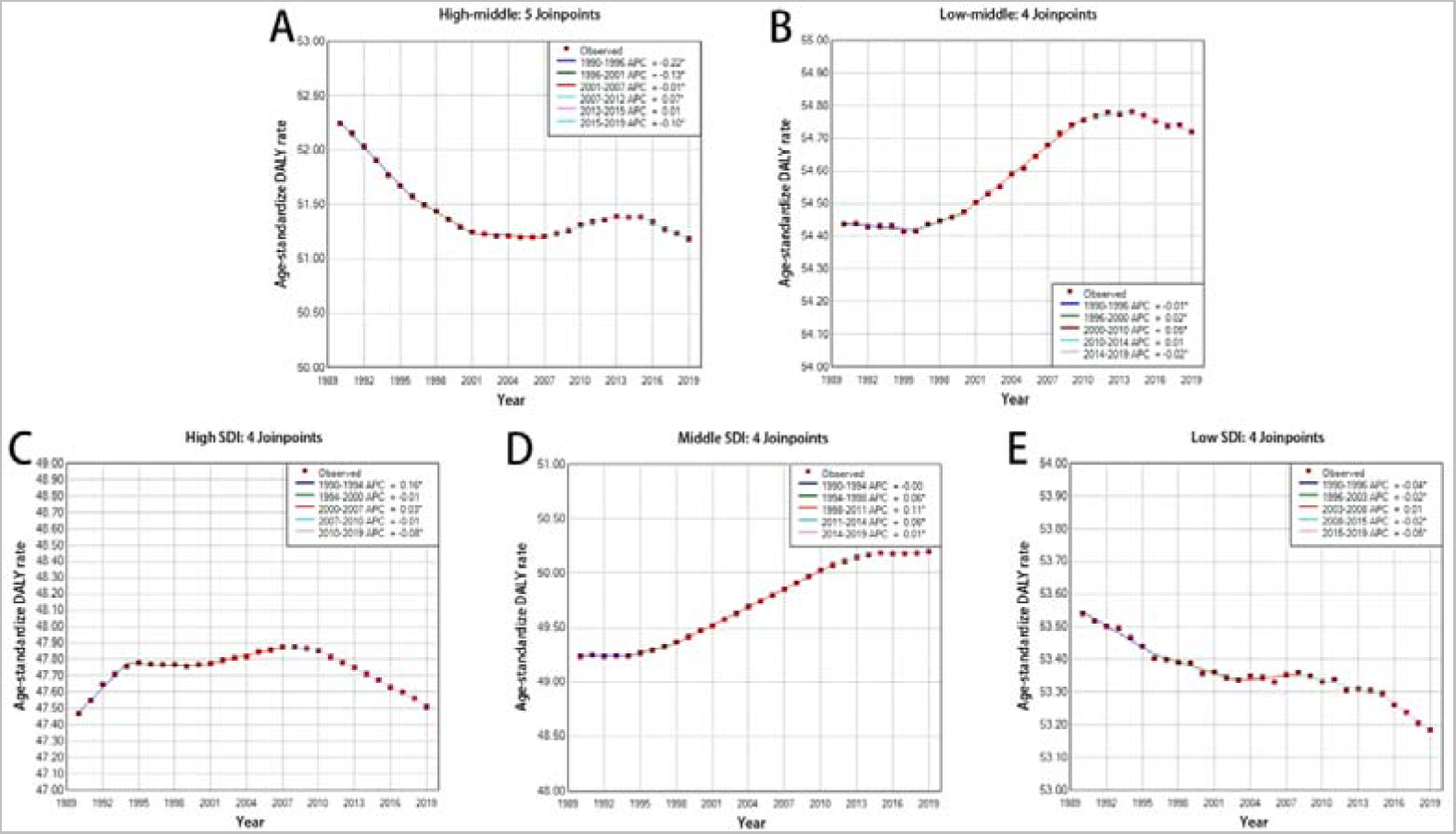
Temporal trends of age-standardized DALY in 5SDI levels by Jointpoint regression model. (A) High-middle SDI. (B) Low-middle SDI. (C) High SDI. (D) Middle SDI. (E) Low SDI.

## 4 Discussion

This study is the quantitative analysis of the global burden and trend of urticaria using the Global Burden of Disease (GBD) 2019 database. Through a comprehensive assessment of urticaria’s prevalence, incidence, and DALY of urticaria, this research compares and analyzes data across different countries, regions, age groups, genders, and Social Development Index (SDI). The Global urticaria metrics including ASIR, ASPR, and age-standardized DALYs for urticaria show an increase trend, which is basically similar to the results of previous studies.(16,17) It’s difficult to attribute the cause of urticaria because of its multifactorial etiology. Nonetheless, it can be speculated that the increase of burden is closely related to the improvement in diagnostic levels and detection rates. There are also observational studies have found an increase in allergic diseases due to increased temperatures. Simultaneously, there are observational studies indicating that the rise in temperature has led to an increase of allergic diseases. (18,19) This change may be due to the alteration in antigen exposure and the decrease in the immune system’s tolerance to antigen specificity caused by the rising temperatures.(20) However, attributing the increased burden directly to climate change is extremely difficult to quantify. Differing from most diseases, the incidence of urticaria is higher than the prevalence worldwide. There are several factors that may explain this. First, urticaria can recur within a single year, increasing its incidence, as each recurrence is counted as a new case. By contrast, prevalence is measured at a specific time and only includes people with active disease.(21,22) Second, the disease course of urticaria is generally brief, with symptoms disappearing within hours or days. As a result, the number of active cases at any time is likely lower than the number of total cases over a year. Third, the mild and self-limiting characteristics can lead to under-reporting. Many patients may not seek treatment or report their condition. Consequently, the actual incidence is likely to be higher than the record, while the prevalence may more accurately represent severe or persistent cases.(23)

Metrics of burden display the heterogeneity across different countries and regions. At the regional level of ASIR, ASPR, and age-standardized DALYs, Central Europe, Eastern Europe, and Central Asia exhibit the highest values, whereas East Asia, Oceania, and Western Europe register the lowest. A significant difference lies in the fact that Central Europe, Eastern Europe, and Central Asia are all inland areas, in contrast to East Asia, Oceania, and Western Europe being near the ocean. This observation is consistent with existing time-series experiments indicating that increase of relative humidity can reduce the number of urticaria outpatient visits.(24,25) Additionally, Central Asia, Central and Eastern Europe often have higher levels of air pollution, and there are observational studies that have demonstrated the correlation between air pollution and allergy symptoms with other skin diseases.(24–26) Among the capitals of nations, those of Central Asian countries are at the top of air pollution rank.(27) Multiple observational studies have confirmed that short-term exposure to air pollutants such as O_3_, PM_2.5_, and PM_10_ increases the risk of urticaria and exhibits a cumulative effect.(18,25) One hypothesis for this phenomenon is that air pollutants induce abnormal oxidative stress and impair the skin barrier.(28,29) We also find that South Asia has the largest growth trends in ASIR, ASPR, and DALYs. This trend can attribute to the rapid economic development represented by countries like India and Bangladesh. Such development is accompanied by swift population growth and advances in local medical technology, resulting in increased outpatient visits. Moreover, industrial growth contributes to pollution, thereby exacerbating both the incidence and prevalence of diseases in the South Asia region.(27)

At the national level, Nepal obtains the highest values in ASIR, ASPR, and age-standardized DALYs. We believe that the factors for such a phenomenon are multifactorial. Geographically located on the southern side of the Himalayas with an average elevation of 3,265 meters, Nepal’s high-altitude areas are subjected to strong ultraviolet rays, which is are more likely to damage the skin barrier, consequently instigating the incidence of urticaria.(30,31) Concurrently, the abundant vegetation in the southern Himalayan produces amounts of pollen, a typical allergen, serving as a common trigger for urticaria. The abundant vegetation also attracts a substantial insect population, commonly leading contact urticaria.(32) Considering Nepal’s traditional culture, the use of spices may also be a potential risk factor, though there is no experimental evidence has yet to confirm this. Along with a wealth of disease-causative factors and low per capita healthcare expenditure, Nepal ranks highest in ASIR, ASPR, and DALYs.

In terms of gender, the females’ burden is higher than the males’ burden. In all age groups, females exhibit higher values of ASIR, ASPR, and age-standardized DALYs than males, resembling previous research findings.(5,9,16,17,33) In terms of age, the incidence, prevalence, and DALYs for the <14 years age group significantly outweigh those for the >14 years age group, confirming prior studies as well.(25,17,34) The high ASIR and ASPR in females are speculated to be influenced by estrogen, as animal studies have proved its capacity to promote mast cell activation and allergic sensitization(35). Additionally, there is research shows that air pollution is more likely to harm female skin.(26)

In terms of the relationship between the Socio-Demographic Index (SDI) and the burden of urticaria, there is a positive correlation between SDI and the burden of urticaria. In the 6SDI statistics, it is found that low-middle SDI and middle SDI groups have higher burden in comparison to other groups. Additionally, children <14 years old had the highest proportion of all cases. Individual and family income are recognized as significant determinants of health. There are studies similar to ours indicate that children from households with higher incomes face higher risks of urticaria compared to children from lower-income households.(36) But there also are experiments that, contrary to our conclusions, demonstrating that individuals from regions with a higher poverty percentage exhibit a higher risk of urticaria visits.(37) For such differences, several reasons are posited: Firstly, reporting bias: regions with high SDI often have advanced healthcare systems, potentially increasing the diagnostic rates for urticaria, thereby increasing prevalence and incidence rates. In regions with low SDI, many cases may remain unreported or undiagnosed due to inadequate diagnostic facilities or lack of public awareness. Secondly, lifestyle factors: regions with high SDI, high living standards and levels of urbanization are more likely to touch environmental allergens, medications and food additives, which are potential triggers for urticaria.(38) Thirdly, experimental differences: existing observational studies focus on some individual subjects and are somewhat randomized, whereas the Global Burden of Disease (GBD) study focuses on regional group and is more representative.

In terms of the development of the burden of urticaria, the jointpoint graphs show that the middle SDI region has the most significant increase in all burden indicators from 1990-2019. The special features of middle SDI regions are posited to be contributory factors. These regions are likely undergoing processes of industrialization and urbanization, increasing environmental and lifestyle-related disease triggers, which leads to an increase in both the incidence and prevalence of urticaria.(18,38) At the same time, middle SDI regions may be facing additional socio-economic pressures, such as job stress and accelerated life rhythms, which could potentially lead to the release of neurotrophic factors, thereby promoting the activation of mast cells in the skin and resulting in the onset of urticaria.(39–41)

This study provides a comprehensive survey of the global, national, and regional burden of urticaria. Within the specific disease category of urticaria, there are several different subtypes, including but not limited to acute versus chronic urticaria and spontaneous versus induced urticaria, and GBD studies often fail to make a clear distinction between these subtypes due to limitations in statistical techniques or data availability, leading to difficulties in attributing prevalence and burden, which in turn impacts the relevance of disease prevention and control policies. Second, GBD may utilize preferred case definitions or alternative measures when processing data, which is particularly evident when data are scarce or of low quality. In studies of urticaria, the possibility exists that prevalence and incidence are underestimated because cases with mild symptoms may not be reported or treated, and such underestimates can be misleading about the distribution of the disease burden. Overall, the limitations of the GBD study on urticaria are mainly in the availability, accuracy, and clarity of data for specific disease classifications, which may have implications for disease burden estimates and corresponding public health policies.

## 5 Conclusion

This study indicates that from 1990 to 2019, the global burden of urticaria has been on an upward trend, showing obvious geographical heterogeneity, thereby proving the necessity for region-specific interventions. In terms of gender, females get a higher disease burden than males. Concurrently, a correlation exists between the Social Demographic Index (SDI) and the burden of urticaria. These findings remind us of the need for further epidemiological research and targeted public health policies to more accurately interpret etiological factors and reduce the disease burden.

## Data Availability

All data produced in the present work are contained in the manuscript.

## Abbreviations

GBD: Global Burden of Disease
DALY: Disability-Adjusted Life Years
YLD: Year lived with disability
EAPC: Estimated Annual Percentage Change
AAPC: Age-Adjusted Percentage Change
SDI: Socio-demographic Index
ASR: Age-Standardized Rate
ASIR: age-standardized incidence rate
ASPR: age-standardized incidence rate

